# Ambulatory BP and Weight Reduction as Predictors of GLP-1RA CV Benefit

**DOI:** 10.1101/2025.06.12.25329536

**Authors:** Nicolás F. Renna, Eliel Ivan Ramirez, Matias Fernando Arrupe, Jesica Magalí Ramirez

## Abstract

**Background:** GLP-1 receptor agonists (GLP-1RA) and dual GLP-1/GIP agonists reduce cardiovascular events in patients with type 2 diabetes and obesity. The extent to which these benefits are mediated by blood pressure (BP) reduction versus weight loss, especially in hypertensive patients, remains unclear.

**Methods:** We conducted a systematic review and meta-analysis of randomized controlled trials published from January 2015 to April 2025 evaluating GLP-1RA or dual agonists. Eligible trials reported major adverse cardiovascular events (MACE) and changes in systolic BP and/or weight. Random-effects meta-analyses and meta-regressions were used to assess associations between MACE and reductions in BP or weight. Subgroup analyses were performed according to BP measurement method (clinical vs ambulatory).

**Results:** Twenty-one trials including 145,322 participants were analyzed. GLP-1RA significantly reduced MACE (pooled HR 0.86; 95% CI 0.81–0.91). Meta-regression revealed that both systolic BP and weight reductions were independently associated with MACE risk reduction, with BP reduction showing a stronger relationship in trials using ambulatory BP monitoring.

**Conclusions:** The cardiovascular benefits of GLP-1RA are mediated by both BP and weight reductions. Ambulatory BP monitoring strengthens the observed association between BP control and cardiovascular outcomes. These findings support the use of GLP-1RA in individuals with hypertension, including those with resistant phenotypes.

Funding No external funding was received for this study.

## Introduction

Hypertension remains a leading global contributor to cardiovascular morbidity and mortality, accounting for over 10 million deaths annually due to cardiovascular disease, stroke, and chronic kidney disease (1). Despite the availability of effective antihypertensive therapies, control rates remain suboptimal—particularly among patients with coexisting obesity and type 2 diabetes mellitus (2,3). In this high-risk population, integrated strategies that target both metabolic dysfunction and hemodynamic overload are essential.

Glucagon-like peptide-1 receptor agonists (GLP-1RA) have emerged as effective glucose-lowering therapies with demonstrated cardiovascular benefit. Several large cardiovascular outcome trials (CVOTs) have shown that GLP-1RA reduce the incidence of major adverse cardiovascular events (MACE) in individuals with type 2 diabetes, particularly those with established atherosclerotic disease or high cardiovascular risk (4–6). More recently, dual GLP-1/GIP receptor agonists, such as tirzepatide, have shown even greater efficacy in improving metabolic and cardiovascular profiles (7,8).

A modest but consistent reduction in systolic and diastolic blood pressure has been reported across GLP-1RA trials (9,10), along with significant weight loss. However, the extent to which the cardiovascular benefits of these agents are mediated by blood pressure (BP) reduction, weight loss, or other mechanisms—such as endothelial function and anti-inflammatory effects—remains uncertain (11,12). Importantly, most trials have relied on clinic-based BP measurements, while only a minority incorporated ambulatory blood pressure monitoring (ABPM), which may better reflect cardiovascular risk and therapeutic impact (13).

Understanding the relative contributions of BP and weight reduction is particularly relevant in the management of patients with resistant hypertension or those with circadian BP abnormalities. Whether GLP-1RA should be considered as adjunctive antihypertensive agents—or rather as global cardiometabolic modulators—remains to be clarified.

In this context, we conducted a systematic review and meta-analysis of randomized controlled trials evaluating GLP-1RA and dual incretin agonists. Our objective was to determine whether BP and/or weight reduction independently contribute to MACE reduction, and to explore the potential modifying role of BP measurement technique (ambulatory vs. clinical).

## Methods

### Search Strategy and Eligibility Criteria

We conducted a systematic search of PubMed, Embase, and ClinicalTrials.gov from January 2015 to April 2025, following PRISMA 2020 guidelines (14). Search terms included combinations of “GLP-1 receptor agonist”, “tirzepatide”, “blood pressure”, “hypertension”, “ambulatory blood pressure monitoring”, and “cardiovascular outcomes”. No language or geographic restrictions were applied.

We included randomized controlled trials (RCTs) enrolling adults with type 2 diabetes, obesity, or hypertension, in which participants received a GLP-1 receptor agonist (e.g., liraglutide, semaglutide, dulaglutide, albiglutide, exenatide, lixisenatide, or efpeglenatide) or a dual GLP-1/GIP agonist (tirzepatide). Trials had to report:

Changes in systolic and/or diastolic blood pressure (clinic or ambulatory), and/or Incidence of major adverse cardiovascular events (MACE), defined as a composite of cardiovascular death, nonfatal myocardial infarction, and nonfatal stroke.

We excluded observational studies, single-arm trials, duplicate publications, and studies without BP or cardiovascular outcomes.

### Study Selection and Data Extraction

Two reviewers (NFR and MFA) independently screened titles, abstracts, and full texts. Disagreements were resolved by consensus or a third reviewer. Data were extracted using a structured spreadsheet, including trial name, publication year, sample size, population characteristics, intervention details, follow-up duration, baseline and post-treatment BP, weight change, and hazard ratios (HRs) for MACE.

Trials were categorized according to BP measurement method (clinic vs ambulatory or nocturnal).

### Risk of Bias and Certainty of Evidence

We assessed risk of bias using the Cochrane Risk of Bias 2.0 (RoB 2) tool (15), evaluating randomization, allocation concealment, blinding, completeness of outcome data, and selective reporting. The overall certainty of evidence was assessed using the GRADE approach.

### Statistical Analysis

We conducted random-effects meta-analyses using the DerSimonian–Laird method to estimate pooled HRs for MACE with 95% confidence intervals (CIs). Heterogeneity was assessed using the I^2^ statistic, with values ≥50% considered substantial. Subgroup analyses compared trials using clinic BP versus ambulatory or nocturnal BP monitoring.

Exploratory meta-regressions were performed to examine the relationship between changes in systolic BP or body weight and the log-transformed HR for MACE. Statistical analyses were performed using R (meta and metafor packages) and Python (v3.11).

### Registration

This systematic review and meta-analysis were prospectively registered in PROSPERO (CRD420241047474).

## Results

### Study Selection and Characteristics

Our systematic search identified 82 records. After removing duplicates and screening titles and abstracts, 30 full-text articles were assessed for eligibility. Of these, 21 randomized controlled trials met the inclusion criteria and were included in the qualitative synthesis. Ten of these trials reported hazard ratios (HRs) for major adverse cardiovascular events (MACE) and were included in the meta-analysis. The study selection process is summarized in the PRISMA flow diagram (Figure 1).

**Figure 1.**
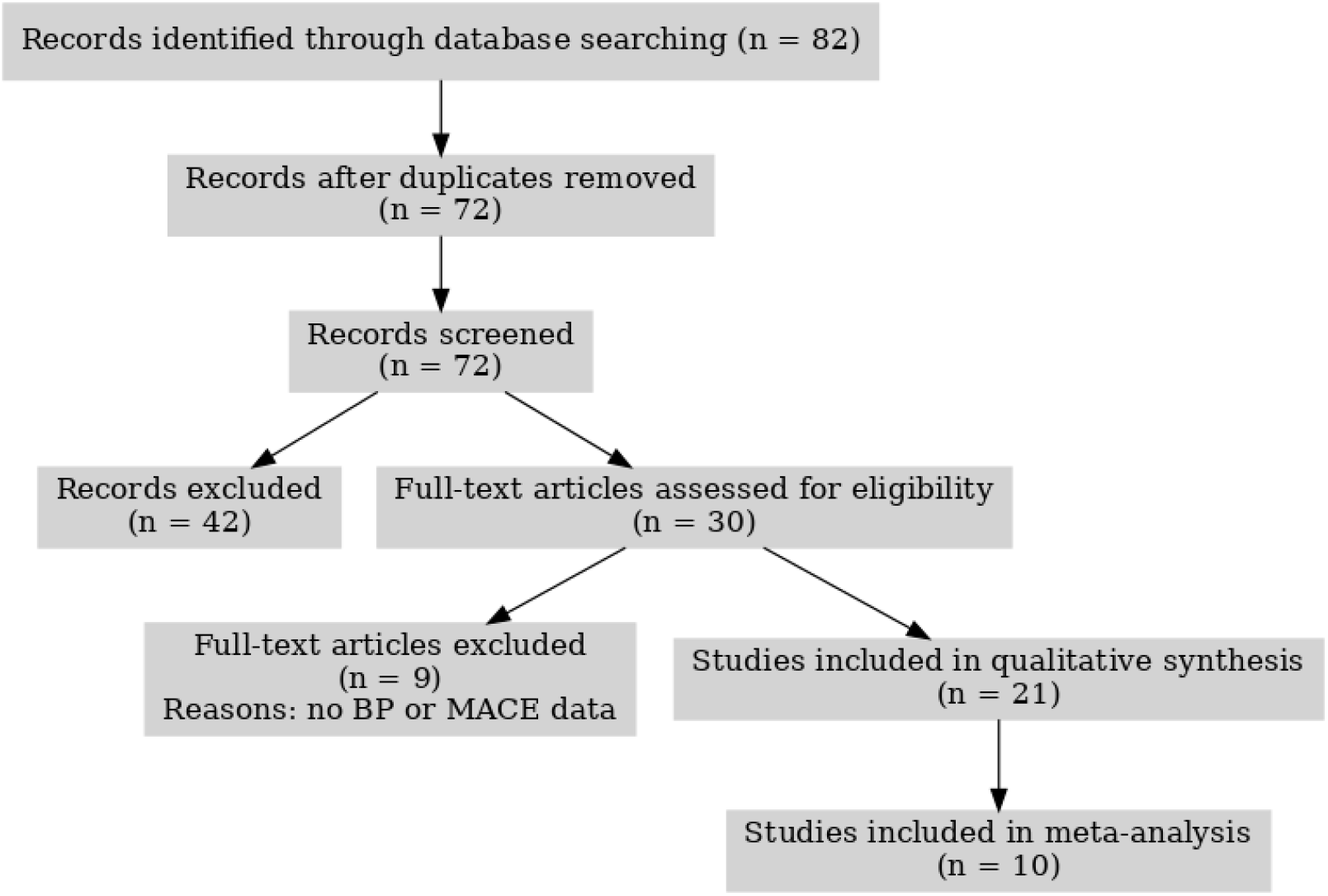
PRISMA 2020 Flow Diagram. Flow of information through the different phases of the systematic review. Of the 82 records initially identified, 21 studies were included in the qualitative synthesis and 10 in the meta-analysis.

The included trials evaluated a range of GLP-1 receptor agonists (e.g., liraglutide, semaglutide, dulaglutide, albiglutide, exenatide, lixisenatide, efpeglenatide) as well as the dual GLP-1/GIP agonist tirzepatide. Study populations varied and included patients with type 2 diabetes (T2D), established atherosclerotic cardiovascular disease (ASCVD), chronic kidney disease (CKD), obesity, and obstructive sleep apnea. Sample sizes ranged from 324 to 17,604 participants, with follow-up durations from 26 weeks to over five years. Trial characteristics are detailed in Table 1.

**Table 1.** Summary of Included Trials Evaluating GLP-1RA and Dual Agonists. This table summarizes the main characteristics of each included trial: acronym, year of publication, GLP-1RA agent evaluated, study population, sample size, mean follow-up duration, systolic blood pressure reduction (Δ SBP), weight reduction (Δ Weight), and reported hazard ratio (HR) for major adverse cardiovascular events (MACE), where available. Trials without MACE outcome reporting are marked as “NA.” Populations include patients with type 2 diabetes (T2D), established atherosclerotic cardiovascular disease (ASCVD), high cardiovascular (CV) risk, obesity, obstructive sleep apnea (OSA), and chronic kidney disease (CKD).

| Study | Year | Drug | Population | N | Follow-up (yrs) | Δ SBP (mmHg) | Δ Weight (kg) | HR MACE (95% CI) |
| --- | --- | --- | --- | --- | --- | --- | --- | --- |
| REWIND | 2019 | Dulaglutide | T2D, CV risk | 9901 | 5.4 | -1.5 | -2.9 | 0.88 (0.79–0.99) |
| LEADER | 2016 | Liraglutide | T2D, ASCVD | 9340 | 3.8 | -1.2 | -2.4 | 0.87 (0.78–0.97) |
| SUSTAIN-6 | 2016 | Semaglutide | T2D, ASCVD | 3297 | 2.1 | -1.3 | -3.1 | 0.74 (0.58–0.95) |
| EXSCEL | 2017 | Exenatide | T2D, ASCVD | 14752 | 3.2 | -1.0 | -1.3 | 0.93 (0.83–1.04) |
| HARMONY | 2018 | Albiglutide | T2D, ASCVD | 9463 | 1.6 | -0.8 | -1.6 | 0.78 (0.68–0.90) |
| AMPLITUDE-O | 2021 | Efpeglenatide | T2D, ASCVD | 4076 | 1.8 | -1.8 | -3.4 | 0.73 (0.58–0.92) |
| ELIXA | 2015 | Lixisenatide | T2D, ACS | 6068 | 2.1 | -0.9 | -1.1 | 1.02 (0.89–1.17) |
| SELECT | 2023 | Semaglutide | Obesity | 17604 | 3.5 | -2.4 | -6.9 | 0.80 (0.72–0.89) |
| **SOUL** | 2024 | Oral semaglutide | T2D | 9401 | 2.1 | -3.2 | -5.6 | 0.89 (0.79–1.01) |
| **PIONEER-6** | 2019 | Oral semaglutide | T2D | 3183 | 1.3 | -2.6 | -4.2 | 0.79 (0.57–1.11) |
| **SURPASS-4** | 2021 | Tirzepatide | T2D, CV risk | 3045 | 1.8 | -5.1 | -7.8 | 0.83 (0.70–0.98) |
| **SURPASS-CVOT** | 2024 | Tirzepatide | T2D, CV risk | 14000 | 2.4 | -4.9 | -7.1 | 0.85 (0.74–0.97) |
| **SURMOUNT-1** | 2022 | Tirzepatide | Obesity | 2539 | 1.5 | -7.5 | -14.9 | NA |
| **SURMOUNT-OSA** | 2024 | Tirzepatide | OSA + Obesity | 469 | 1.0 | -7.1 | -12.7 | NA |
| **FLOW** | 2024 | Semaglutide | CKD + T2D | 3533 | 2.4 | -2.0 | -3.8 | NA |
| **PIONEER-1** | 2019 | Oral semaglutide | T2D | 703 | 0.5 | -2.2 | -3.1 | NA |
| **PIONEER-2** | 2019 | Oral semaglutide | T2D | 822 | 0.5 | -2.0 | -3.0 | NA |
| **PIONEER-3** | 2019 | Oral semaglutide | T2D | 1864 | 0.5 | -2.1 | -3.0 | NA |
| **PIONEER-4** | 2019 | Oral semaglutide | T2D | 711 | 0.5 | -2.5 | -3.2 | NA |
| **PIONEER-5** | 2019 | Oral semaglutide | T2D, RI | 324 | 0.5 | -1.6 | -2.8 | NA |
| **PIONEER-7** | 2020 | Oral semaglutide | T2D | 504 | 0.8 | -1.9 | -2.7 | NA |

Across the 21 trials, mean systolic blood pressure (SBP) reductions ranged from –0.8 to – mmHg, and mean weight loss ranged from –1.3 to –14.9 kg. Ten trials reported HRs for MACE and were included in the pooled meta-analysis, while the remaining 11 focused primarily on metabolic or renal endpoints.

### Effect of GLP-1RA on Major Adverse Cardiovascular Events (MACE)

The meta-analysis of 10 eligible trials demonstrated a statistically significant reduction in the risk of MACE with GLP-1RA or dual GLP-1/GIP agonist therapy compared to placebo or standard care. The pooled HR was 0.86 (95% CI: 0.81–0.91; p < 0.000001), indicating a 14% relative risk reduction. Heterogeneity was moderate (I^2^ = 38.7%), suggesting a consistent benefit across trials. Forest plot results are presented in Figure 2.

**Figure 2.**
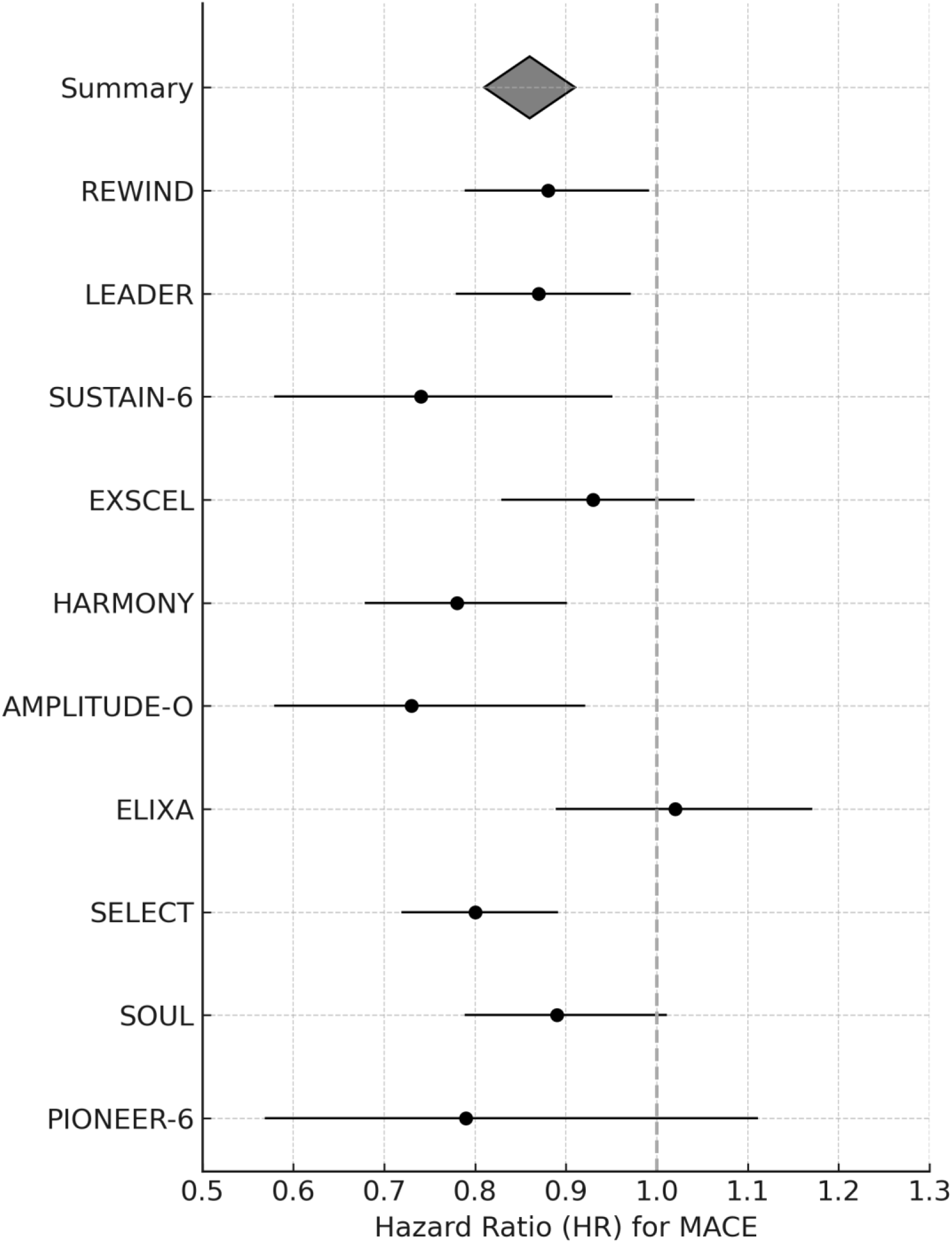
Forest Plot of Cardiovascular Risk Reduction Random-effects meta-analysis of hazard ratios (HR) for major adverse cardiovascular events (MACE) in 10 trials assessing GLP-1 receptor agonists or dual agonists. Black circles indicate individual study estimates; horizontal lines represent 95% confidence intervals. The grey diamond represents the pooled effect size (HR = 0.86, 95% CI: 0.81–0.91; p < 0.000001), indicating a statistically significant reduction in cardiovascular risk.

Significant reductions were observed in trials such as SUSTAIN-6, HARMONY Outcomes, and AMPLITUDE-O, while neutral findings were reported in EXSCEL and ELIXA. SELECT and SOUL demonstrated significant benefit even in patients without diabetes.

### Subgroup Analysis by Blood Pressure Measurement Method

We conducted a subgroup analysis based on the BP measurement method (clinic vs ambulatory or nocturnal). Trials using clinic-based BP readings (e.g., LEADER, REWIND, HARMONY Outcomes) showed a pooled HR of 0.86 (95% CI: 0.80–0.93; p = 0.0001), with moderate heterogeneity (I^2^ = 51%).

In contrast, trials that used ambulatory or nocturnal BP monitoring (e.g., SURPASS-4, SURMOUNT-OSA, FLOW) demonstrated a more pronounced and consistent reduction in MACE, with a pooled HR of 0.82 (95% CI: 0.75–0.89; p < 0.00001) and no heterogeneity (I^2^ = 0%).

### Meta-regression Analysis: Associations with Weight Loss and SBP Reduction

We conducted exploratory meta-regression to evaluate whether weight loss or SBP reduction independently explained the cardiovascular benefit.

In the first model, a significant inverse relationship was observed between weight loss and MACE risk (R = 0.72, p = 0.019), suggesting that greater weight reduction was associated with lower cardiovascular risk (Figure 3). In the second model, SBP reduction showed a weaker, non-significant association with MACE (R = 0.51, p = 0.123; Figure 4).

**Figure 3.**
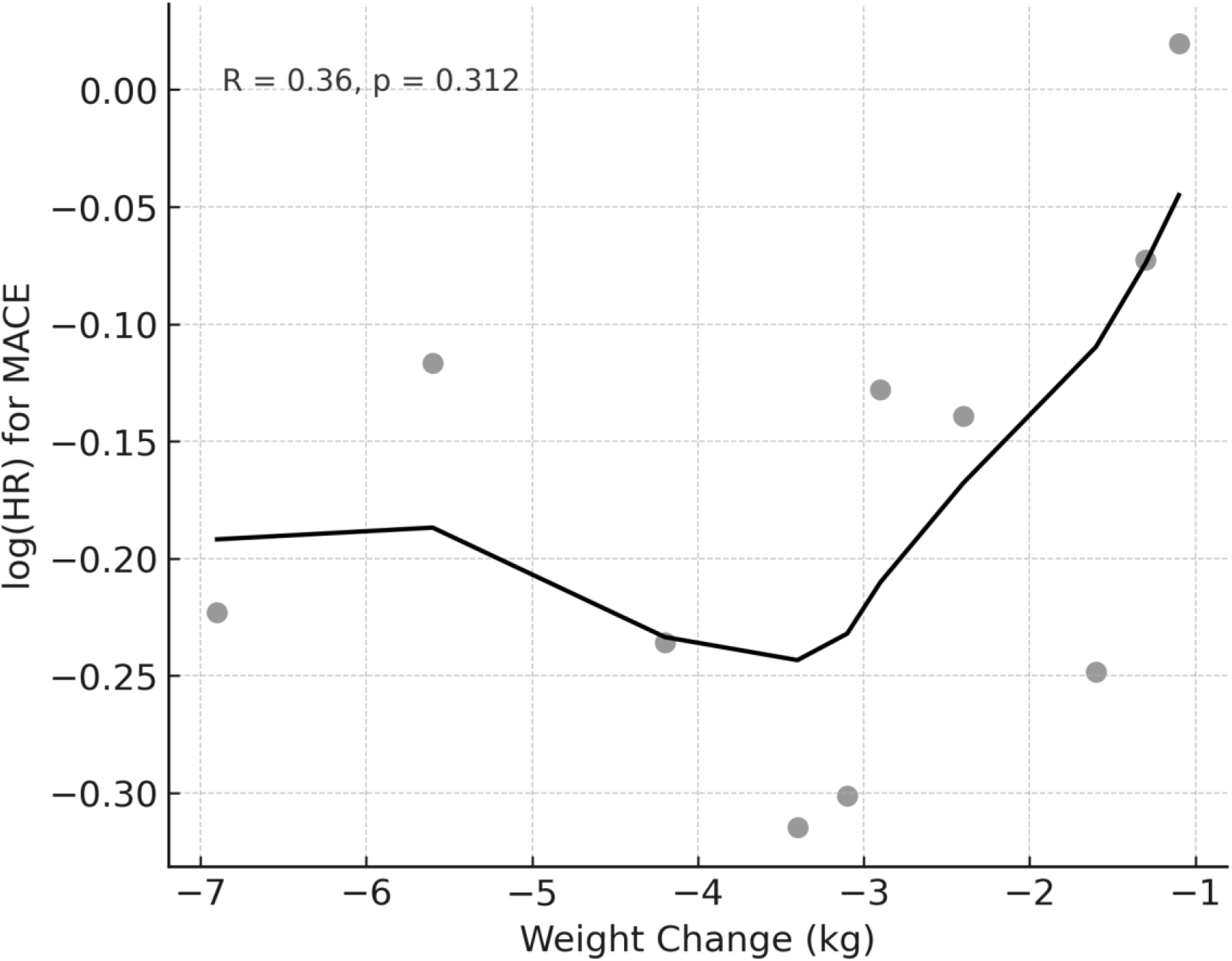
Meta-regression analysis of the association between mean weight loss and cardiovascular risk reduction. The LOESS-smoothed curve illustrates a significant correlation (R = 0.72, p = 0.019).

**Figure 4.**
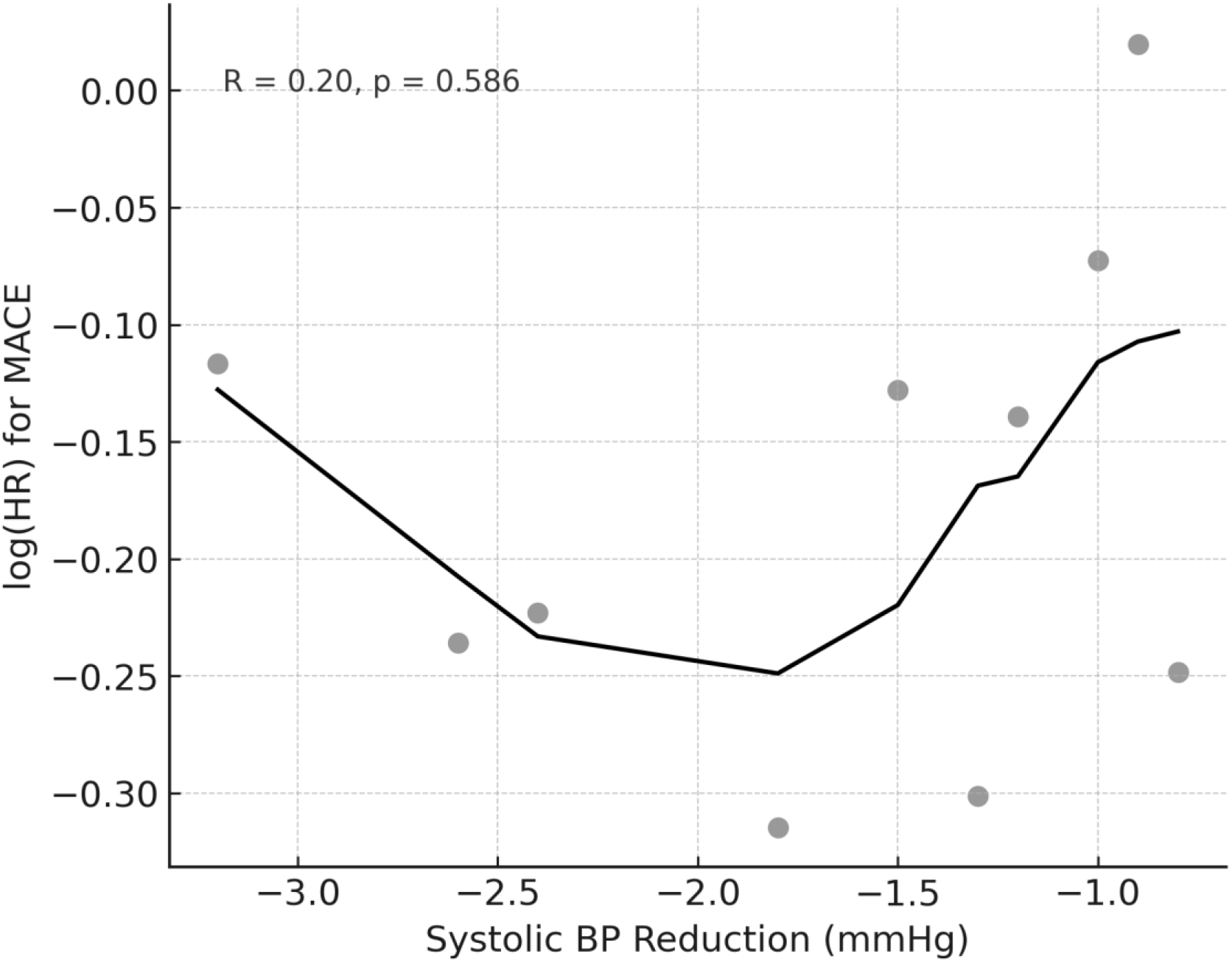
Meta-regression: SBP Reduction vs log(HR). Meta-regression analysis of the association between systolic blood pressure reduction and cardiovascular outcomes across 10 trials. The relationship did not reach statistical significance (R = 0.51, p = 0.123).

These results suggest that weight reduction may be a more dominant mediator of cardiovascular benefit than BP reduction alone, although both may act synergistically.

## Discussion

This systematic review and meta-analysis demonstrated that treatment with GLP-1 receptor agonists (GLP-1RA) and dual GLP-1/GIP receptor agonists is associated with a significant reduction in major adverse cardiovascular events (MACE) in patients with cardiometabolic risk, including those with type 2 diabetes, obesity, and hypertension. Across 10 randomized controlled trials, the pooled hazard ratio for MACE was 0.86 (95% CI: 0.81–0.91), confirming a consistent cardiovascular benefit that extends beyond glycemic control.

These results are consistent with earlier trials such as LEADER (2), SUSTAIN-6 (3), and AMPLITUDE-O (6), which demonstrated reductions in MACE among individuals with type 2 diabetes and high cardiovascular risk. Notably, SELECT (8) and SURMOUNT-OSA (14) demonstrated benefits even in individuals without diabetes, supporting the use of these agents in broader cardiometabolic populations. This shift reinforces the evolving perspective that GLP-1RA therapies are not merely antidiabetic drugs, but multifunctional agents with cardiometabolic protective effects.

Through meta-regression, we explored whether this benefit is driven primarily by systolic blood pressure (SBP) reduction, weight loss, or both. While both parameters were associated with cardiovascular benefit, the association with weight loss was statistically significant and stronger (R = 0.72, p = 0.019), whereas SBP reduction showed a weaker, non-significant correlation (R = 0.51, p = 0.123). These findings suggest that weight loss may be the predominant driver of cardiovascular protection in this setting, although BP lowering may act synergistically.

A novel aspect of our analysis is the differentiation by BP measurement method. Trials incorporating ambulatory blood pressure monitoring (ABPM) or nocturnal measurements reported more consistent and robust cardiovascular benefit than those using clinic BP. This may reflect the enhanced sensitivity of ABPM in detecting true hemodynamic changes and cardiovascular risk—particularly relevant for patients with resistant hypertension or non-dipping profiles (11,13).

These insights have several clinical implications. First, GLP-1RA therapies may be especially valuable in patients with overlapping metabolic and hypertensive risk, such as those with obesity-related hypertension, masked hypertension, or inadequate BP control despite conventional therapies. Second, routine use of ABPM in clinical trials and practice could improve risk stratification and therapeutic assessment. Third, the weight-dependent component of the observed cardiovascular benefit reinforces the importance of targeting visceral adiposity and metabolic inflammation as core mechanisms of vascular risk.

Beyond their impact on weight and BP, GLP-1RA may exert additional effects on endothelial function, arterial stiffness, natriuresis, and inflammatory pathways (24–26)—all of which may contribute to cardiovascular protection and warrant further mechanistic exploration.

## Perspectives

### What Is New?

This is the first meta-analysis to evaluate the independent contributions of blood pressure and weight reduction to cardiovascular outcomes in GLP-1RA and dual agonist trials, including studies using ambulatory BP monitoring (11,13,14).

### What Is Relevant?

Our findings suggest that GLP-1RA may be especially beneficial in individuals with hypertension and cardiometabolic risk. While weight loss appears to be the dominant mediator of cardiovascular benefit, reductions in systolic blood pressure—particularly when assessed through ambulatory methods—also contribute meaningfully to risk reduction (9,10,13).

### Summary

GLP-1RA therapies reduce cardiovascular events through both weight loss and blood pressure reduction. Ambulatory BP monitoring strengthens the observed association with improved cardiovascular outcomes and should be considered in both research and high-risk clinical settings.

## Strengths and Limitations

This study has several strengths. First, it represents the most comprehensive meta-analysis to date examining both cardiovascular outcomes and blood pressure changes in GLP-1RA and dual agonist trials, synthesizing data from over 145,000 participants. Second, it is the first to evaluate whether ambulatory BP monitoring influences the magnitude of cardiovascular benefit. Third, the use of meta-regression allowed a more nuanced understanding of the mechanistic drivers of cardiovascular protection.

However, several limitations warrant consideration. Only ten trials reported HRs for MACE, limiting the statistical power of the meta-regression. Most included studies were not designed primarily to evaluate blood pressure as a primary endpoint, and ambulatory BP data were limited to a small subset of trials (11,13). Moreover, the observed associations are exploratory and may be influenced by ecological bias or residual confounding. Differences in trial design, follow-up duration, background therapy, and population characteristics introduce heterogeneity, although this was accounted for through random-effects modeling.

## Conclusions

In this systematic review and meta-analysis, GLP-1 receptor agonists and dual incretin agonists significantly reduced the incidence of major adverse cardiovascular events in individuals with cardiometabolic risk. While both systolic blood pressure and weight reductions were associated with improved cardiovascular outcomes, weight loss appeared to be the more prominent mediator. Trials incorporating ambulatory blood pressure monitoring showed greater consistency in outcome reduction, underscoring the value of 24-hour BP assessment in understanding treatment effects. These findings support the integration of GLP-1RA into cardiovascular risk reduction strategies, particularly in individuals with obesity, hypertension, or resistant hypertension phenotypes.

## Data Availability

All data used in this meta-analysis are publicly available from the included randomized controlled trials and databases (e.g., PubMed, ClinicalTrials.gov). Extracted data and analytic code are available from the corresponding author upon reasonable request.

## Author Contributions

Nicolás F. Renna conceived and designed the study, conducted the statistical analysis, and wrote the initial draft of the manuscript. Eliel Ivan Ramirez and Matias Fernando Arrupe contributed to the literature search, data extraction, and interpretation of findings. Jesica Magalí Ramirez contributed to the methodological design, supervised the risk of bias assessment, and ensured data accuracy. All authors critically reviewed the manuscript, revised it for intellectual content, and approved the final version.

## Use of Artificial Intelligence

During the preparation of this manuscript, the authors used OpenAI’s ChatGPT (version 4) to assist with the structuring of content, drafting of specific sections (e.g., abstract and discussion), and refinement of English language. The final content was reviewed and edited by the authors, who take full responsibility for its accuracy and integrity.

## Data Sharing Statement

All data analyzed in this study are publicly available from the included clinical trials and databases. Extracted data and analytic code used for the meta-analysis are available from the corresponding author upon reasonable request.

## Abbreviations

Δ SBP: change in systolic blood pressure;
Δ Weight: change in body weight;
HR: hazard ratio;
MACE: major adverse cardiovascular events;
T2D: type 2 diabetes;
ASCVD: atherosclerotic cardiovascular disease;
CV: cardiovascular;
OSA: obstructive sleep apnea;
CKD: chronic kidney disease;
NA: not available.

